# Racial and Ethnic Inequities in Mortality During Hospitalization for Traumatic Brain Injury: A Call to Action

**DOI:** 10.1101/2021.04.06.21254905

**Authors:** Emma A. Richie, Joseph G. Nugent, Ahmed M. Raslan

**Author notes:** **Correspondence:** Emma A. Richie.

## Abstract

The health disparities which drive inequities in health outcomes have long plagued our already worn healthcare system and are often dismissed as being a result of social determinants of health. Herein, we explore the nature of these inequities by comparing outcomes for racial and ethnic minorities patients suffering from traumatic brain injury (TBI). We retrospectively reviewed all patients enrolled in the Trauma One Database at the Oregon Health & Science University Hospital from 2006 to October 2017 with an abbreviated injury scale (AIS) for the head or neck greater than 2. Racial and ethnic minority patients were defined as non-White or Hispanic. A total of 6,352 patients were included in our analysis with 1,504 in the racial and ethnic minority cohort vs. 4,848 in the non-minority cohort. A propensity score (PS) model was generated to account for differences in baseline characteristics between these cohorts to generate 1,500 matched pairs. The adjusted hazard ratio for in-hospital mortality for minority patients was 2.21 (95% Confidence Interval (CI) 1.43-3.41, p<0.001) using injury type, probability of survival, and operative status as covariates. Overall, this study is the first to specifically look at racial and ethnic disparities in the field of neurosurgical trauma. This research has demonstrated significant inequities in the mortality of TBI patients based on race and ethnicity and indicates a substantive need to reshape the current healthcare system and advocate for safer and more supportive pre-hospital social systems to prevent these life-threatening sequelae.

## 1 Introduction

Structural disparities propagated against racial and ethnic minorities have long been known to drive inequities in health outcomes in the US yet are often dismissed as an unfortunate yet unavoidable consequences of what has been disguised as the social determinants of health. However, many of these inequities are formed long before our patients ever step foot into a healthcare facility and stem not from individual poor health decisions but from decades of structural racism and systematic oppression which leave minority patients at a disadvantage for health outcomes even from before birth^(1-3)^. (It is important to state clearly at this point that we will discuss race and ethnicity not as biologically-based separations of persons but as socially-formed systematic manners of categorizing persons based on culture, geographic origin, language, etc. so as to provide financial, social, and political advantage over specific groups of persons with a long and evidence-based history of such action^(3-8)^.) These systemic factors have been extensively researched in the fields of public health, sociology, psychology, and more to demonstrate significant negative impacts on a minority persons lifelong risk for medical and psychological trauma^(1-3,9)^.

Specifically in the field of trauma, these inequities are then exacerbated by ongoing external factors such as insurance, access to level I trauma centers, mechanism of injury, time to surgical intervention, staffing of skilled providers, access to post-acute care and rehabilitation, and community or financial support^(1, 10-14)^. In neurosurgical trauma, the crux of disease burden lies in traumatic brain injury (TBI) which is increasingly becoming a more prevalent global medical issue with devastating effects on both personal and healthcare sequelae^(15-20)^. As with other healthcare fields, neurosurgery and neuro-trauma are not immune to the adverse effects of structural racism on racial and ethnic inequities^(10, 21-24)^. Misguided past research in this field has failed to identify or call out the specific role of sociological and systemic factors that influence these pervasive, persistent, and problematic disparities of health outcomes for our minority patients. In this study, we explore the racial and ethnic inequities in mortality during hospitalization following traumatic brain injury and will engage both a sociological and medical perspective to call to action a change in practice towards a more expansive and sympathetic understanding of the factors involved in minority health outcomes in neuro-trauma.

## 2 Materials and Methods

### 2.1 Study design and setting

This study is a retrospective review of all patients entered into the Trauma One Database established in 2006 at the Oregon Health & Science University Hospital (OHSU), one of the two Level I Trauma centers in the state of Oregon. For our research, we included all patients ≥ 16 years-old entered into the system from 2006 to October of 2017. The dataset used for analysis contained all patient demographic and insurance characteristics, details regarding the mechanism of injury (MOI), treatments provided in the pre-hospital setting and throughout the hospital stay, comorbidities at the time of presentation, discharge disposition, and overall complications. This study follows the Strengthening the Reporting of Observational Studies in Epidemiology (STROBE) reporting guidelines, and was approved by the OHSU Institutional Review Board^(25)^.

### 2.2 Measurements and Outcomes

TBI severity was classified according to the patient’s Glasgow Coma Scale (GCS) score on arrival with the following classifications: severe as GCS <8, moderate as 8-12, and mild as ≥ 13. Minor trauma was defined by an Injury Severity Score (ISS) of ≤ 15. Racial and ethnic minority patients were defined as identifying as any non-White race or as being of Hispanic ethnicity. Mechanism of injury categories were assigned based on the Abbreviated Injury Scale (AIS), ISS, and ICD-9-CM, or ICD-10-CM codes^(26)^. Probability of survival is calculated at our institution using the Trauma Injury Severity Score methodology with a weighted formula comprising of ISS, age, and Revised Trauma Score.

The primary outcome of interest was in-hospital mortality stratified by racial and ethnic minority status. The secondary outcome was the trend in mortality classified by arrival year, injury type, and the presence of work-related injuries.

### 2.3 Statistical Analysis

Continuous variables were described using means and standard deviations (SD), and compared between groups using a two-sample Student’s t-test. Dichotomous and categorical variables were reported using counts and percentages and compared using Fisher’s exact test.

Due to the inherent potential for selection bias in this observational retrospective study, and to account for differences in baseline socioeconomic and demographic characteristics, a propensity-score (PS) matching model was employed. Age, sex, injury mechanism, and primary payor were included as the covariates in this final model, consistent with what has been done in the literature previously when examining racial and ethnic disparities^(27, 28)^. Matching with a 1:1 ratio using nearest neighbor methodology was then performed using MatchIt Package in R, version 4.1.0^(29)^. Balance of baseline covariates and region of common support was then assessed using absolute standardized mean differences, jitter plots and histograms, provided in the supplementary materials.

A multivariate Cox Proportional Hazards regression model was then created using an iterative combined forward and backward stepwise selection procedure from the candidate covariates for the propensity-matched cohorts. Assumptions required for proportional hazards regression were assessed numerically for each covariate, and graphically by the plotting of scaled Schoenfeld residuals. The final model included injury type, probability of survival, and operative status as covariates. Since the AIS score covariate was found to violate the assumption of proportional hazards, the final model was stratified by this covariate. Goodness of fit of each model was then assessed using Cox-Snell residuals.

A p-value < 0.05 was considered to be statistically significant. All analyses were performed using R software, version 4.0.4 (R Foundation for Statistical Computing, Vienna, Austria). The final analysis was confirmed using Stata, version 16.1 (StataCorp, College Station, Texas).

## 3 Results

### 3.1 Patient demographics

There was a total of 6,352 patients included in the Trauma One database with 1,504 included in the racial/ethnic minority cohort and 4,848 patients in the White/non-Hispanic cohort.. The clinical characteristics of the patients and propensity-matched patients are shown in Tables 1A and 1B, respectively. Following statistical PS matching, the maximum number of participants in each group was 1,504 (approximately equivalent to the total number of patients in the minority group).

**Table 1A.**
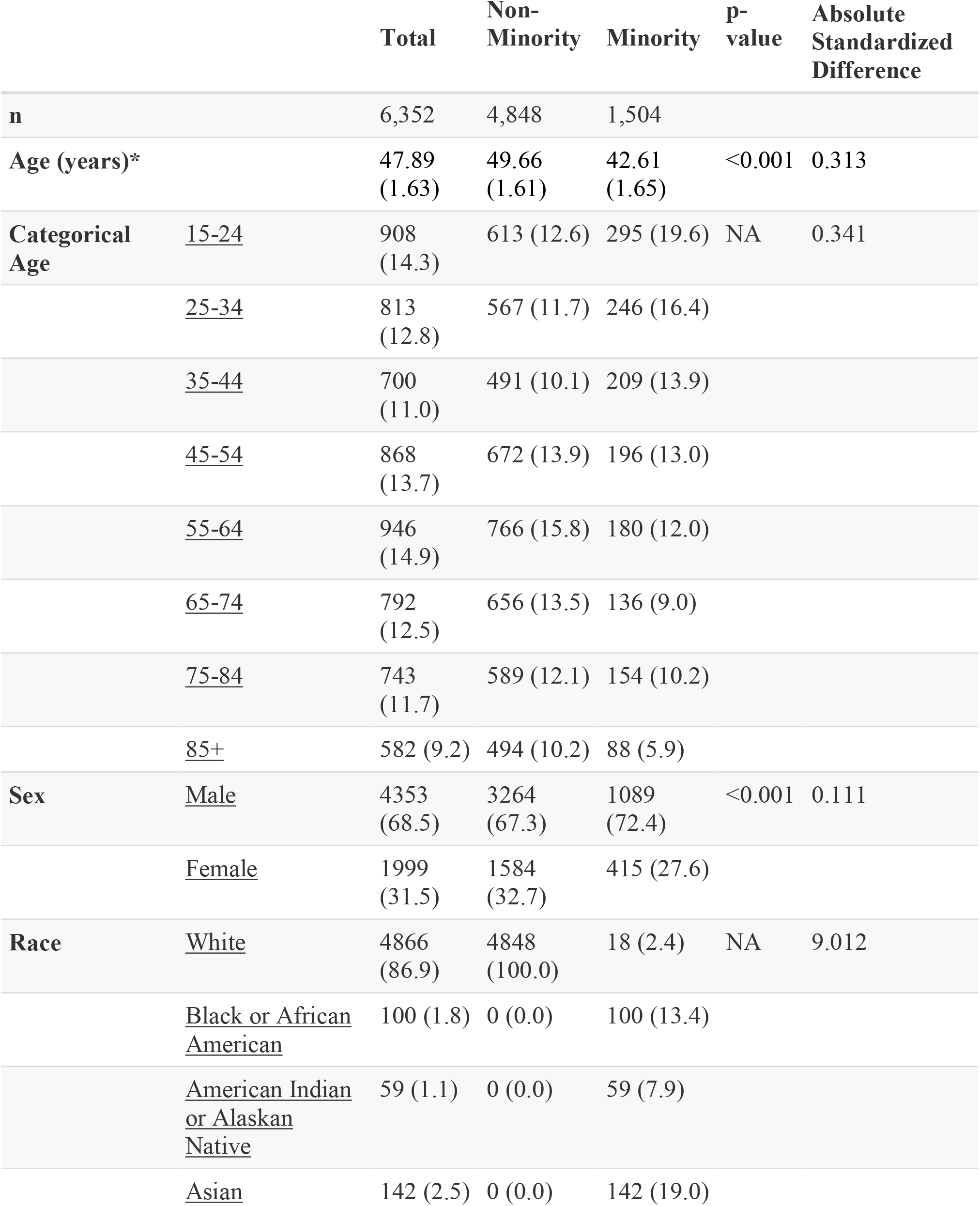
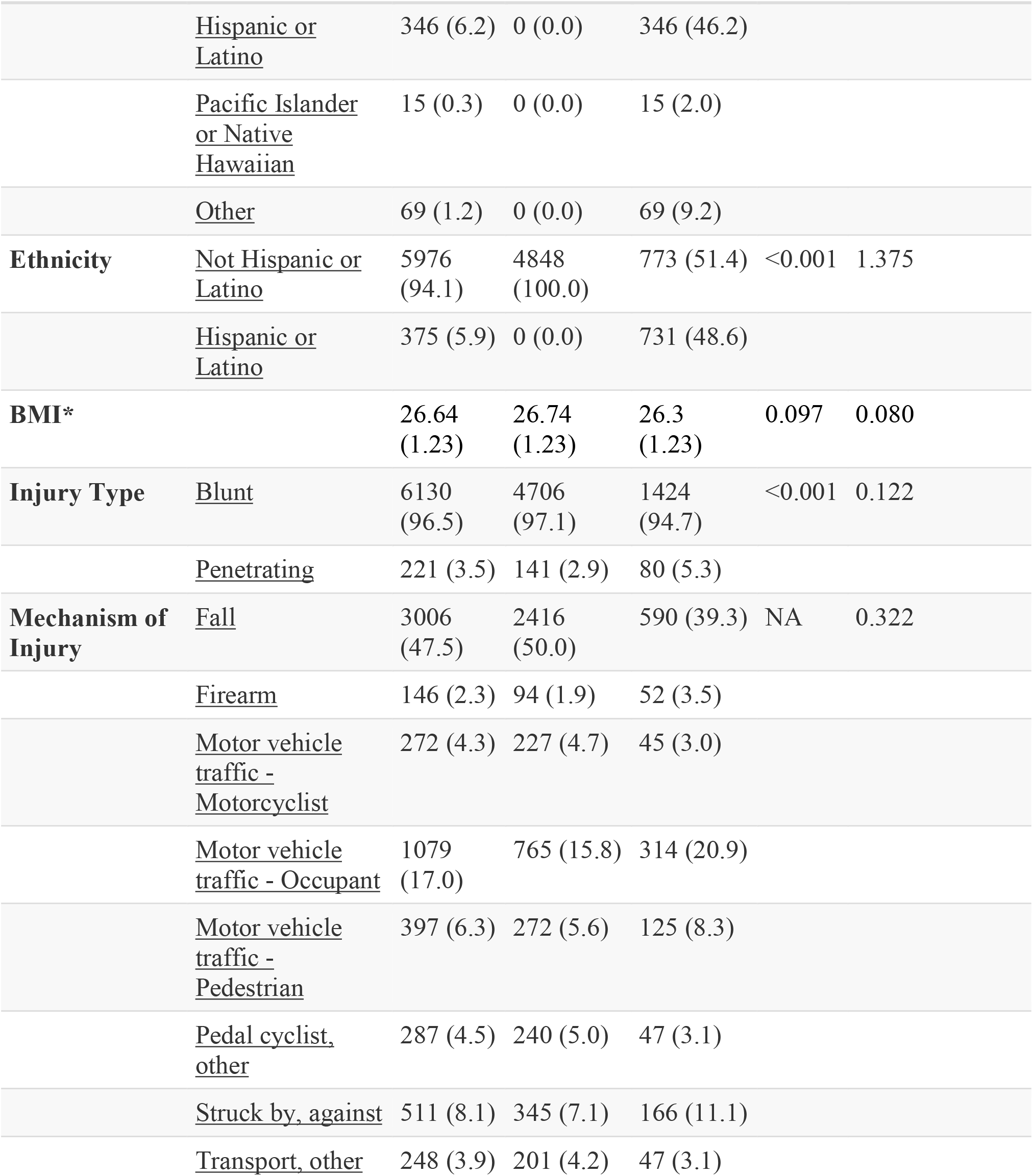
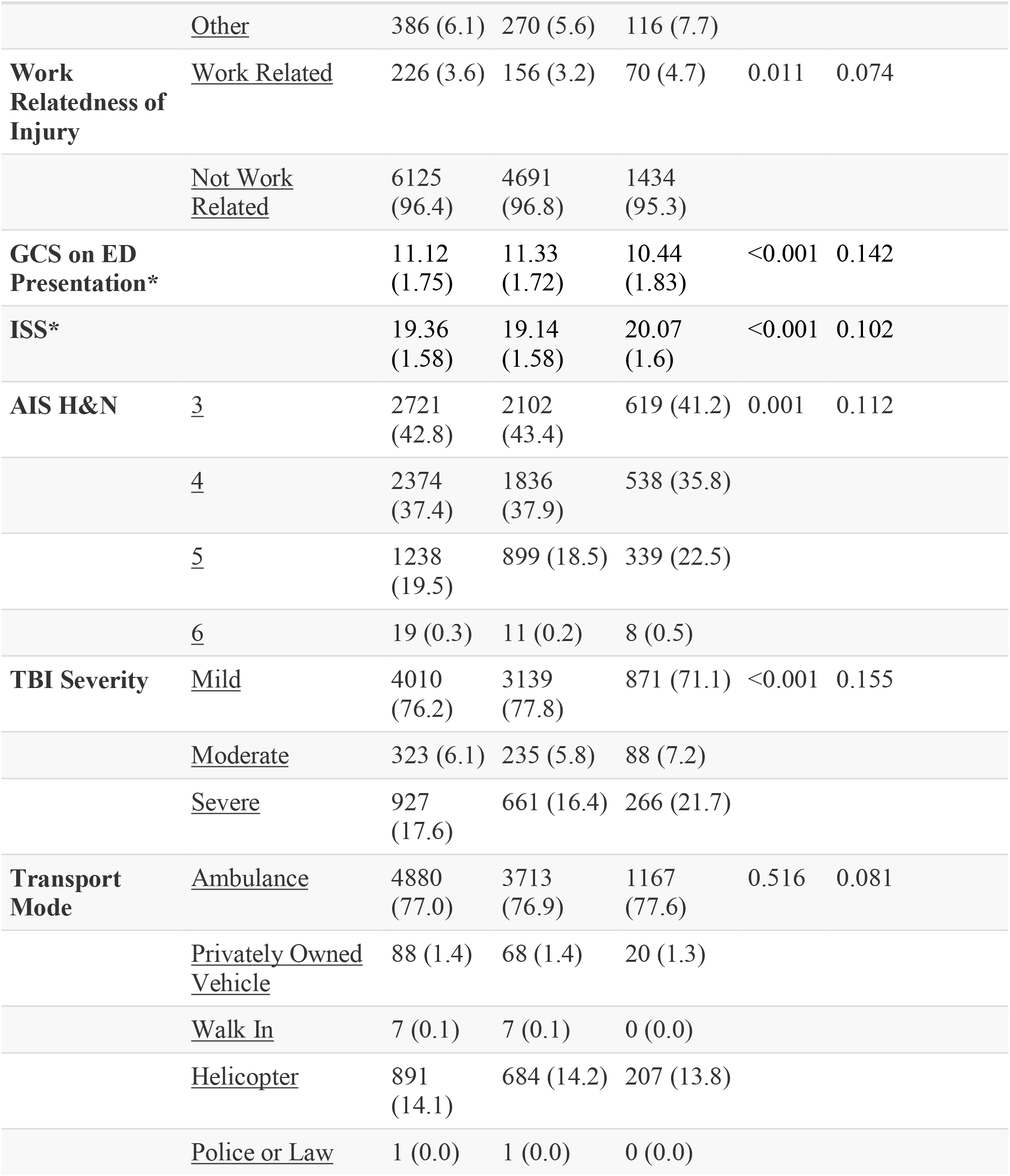
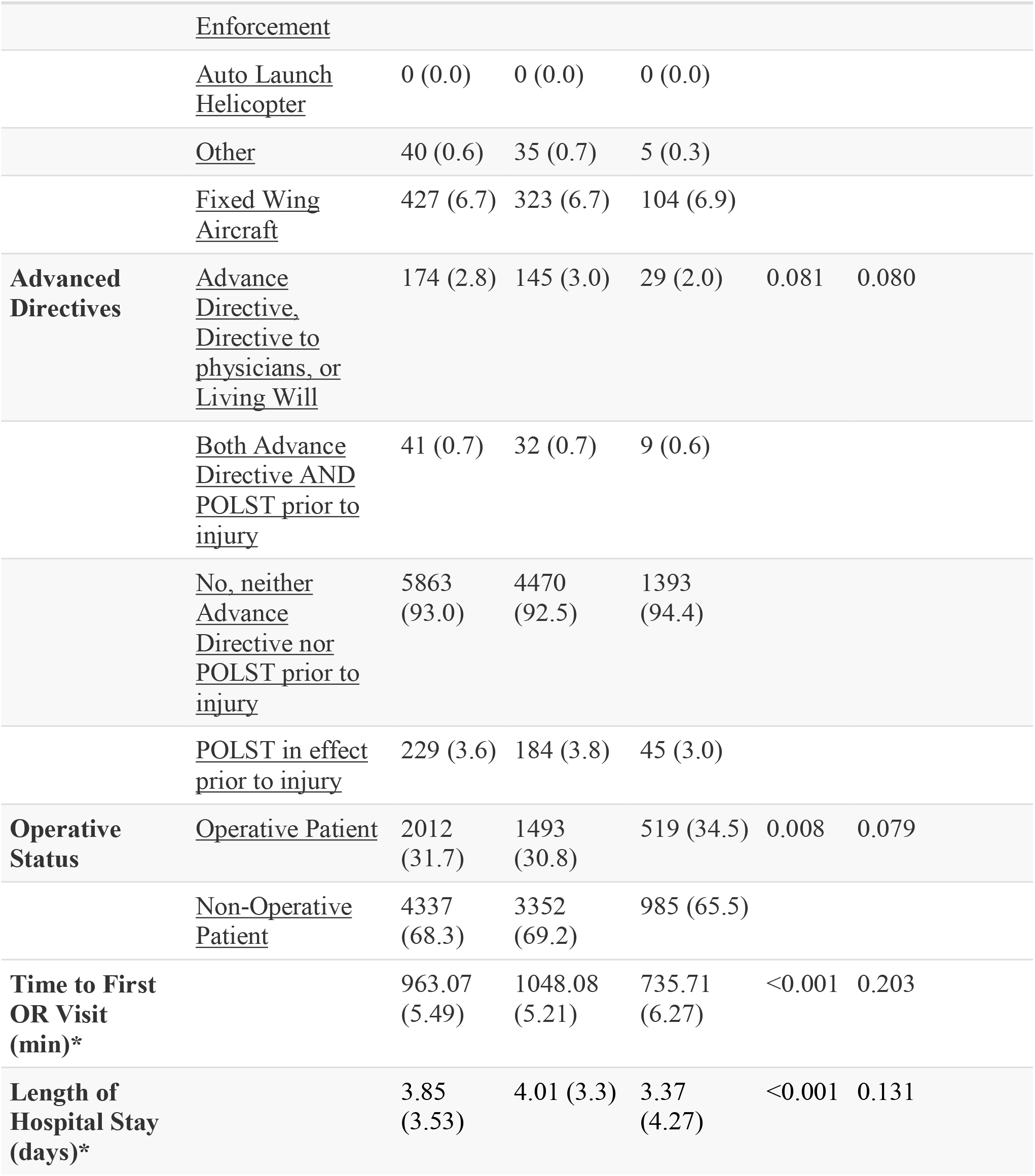
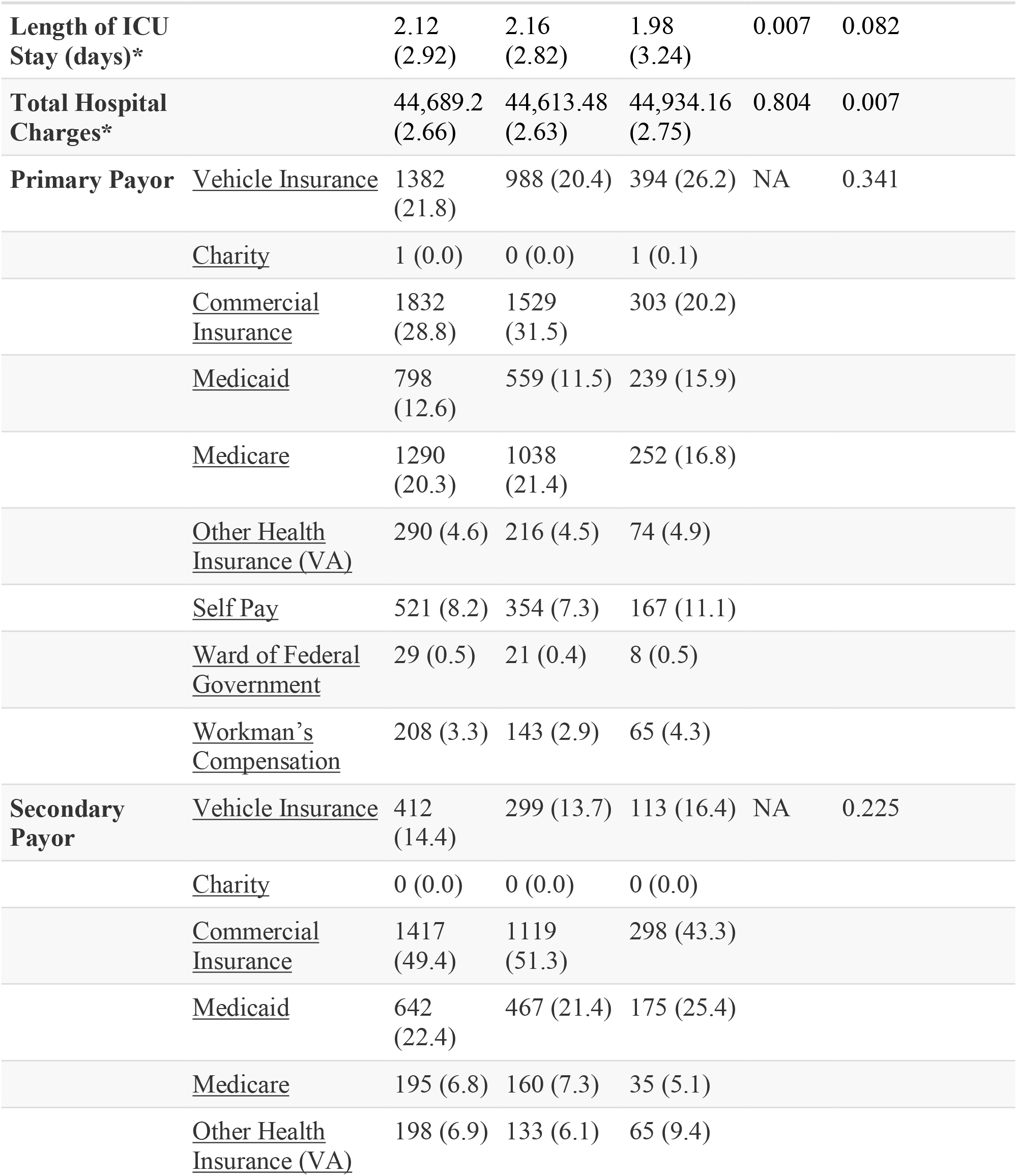
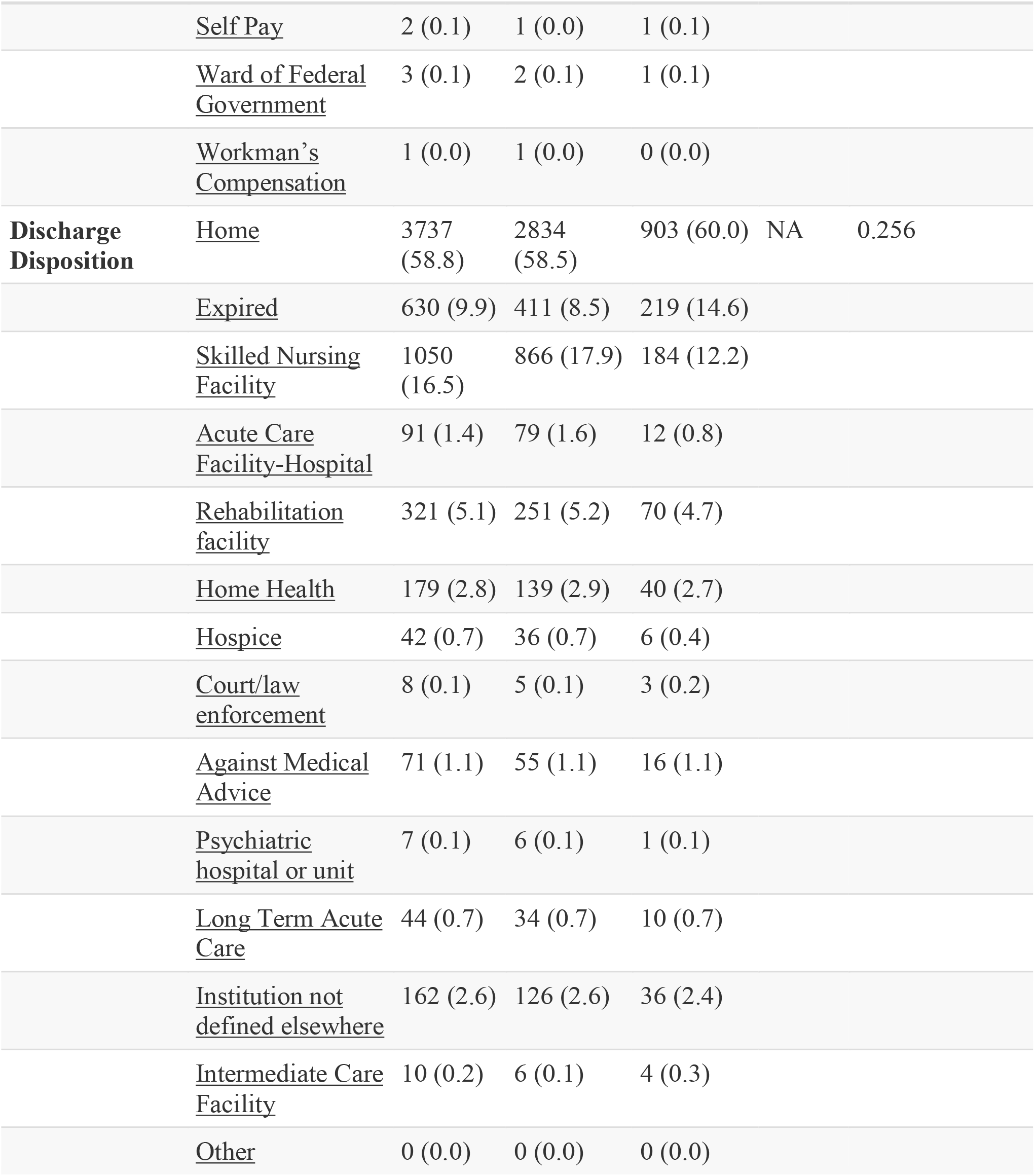
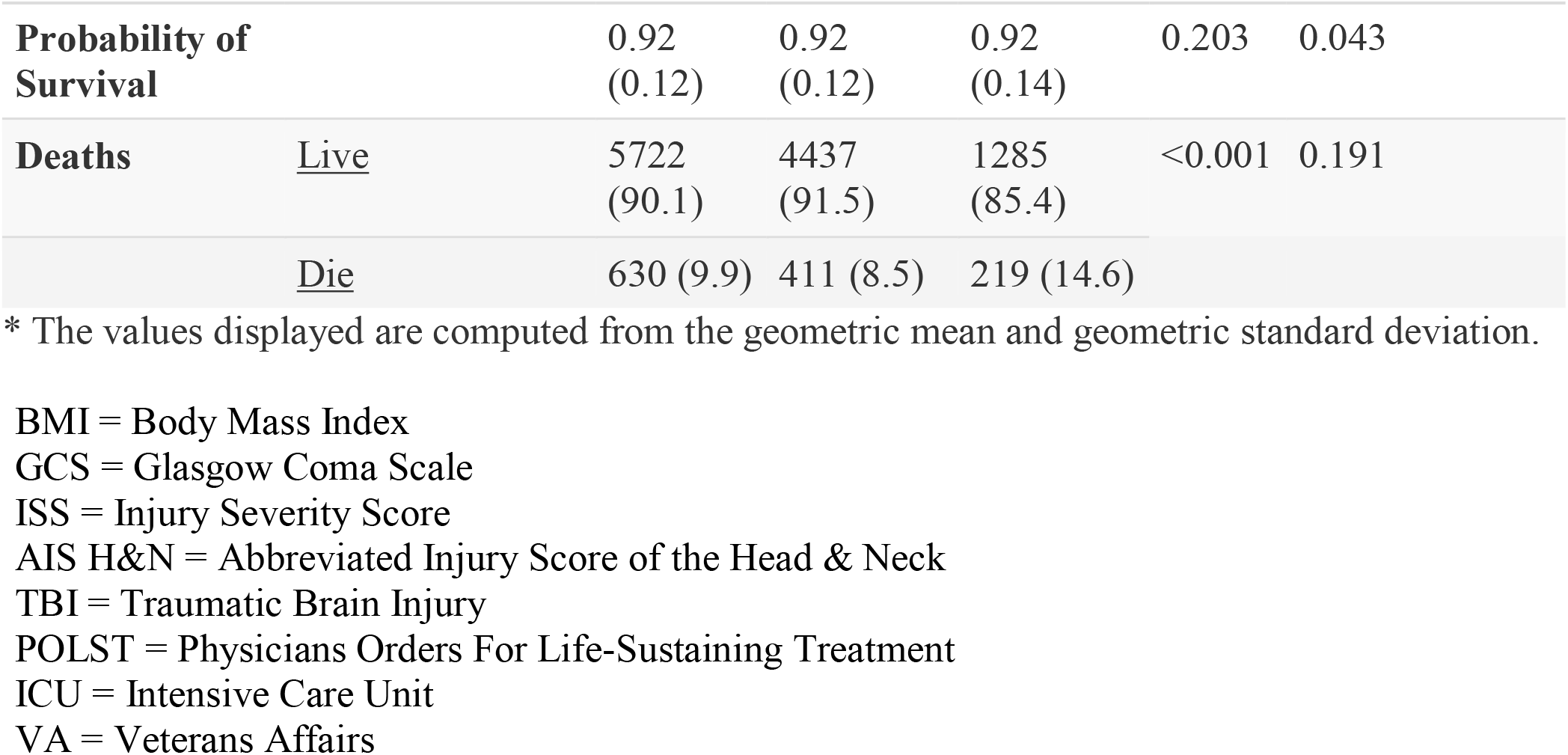
Baseline patient characteristics stratified by minority status.

**Table 1B.**
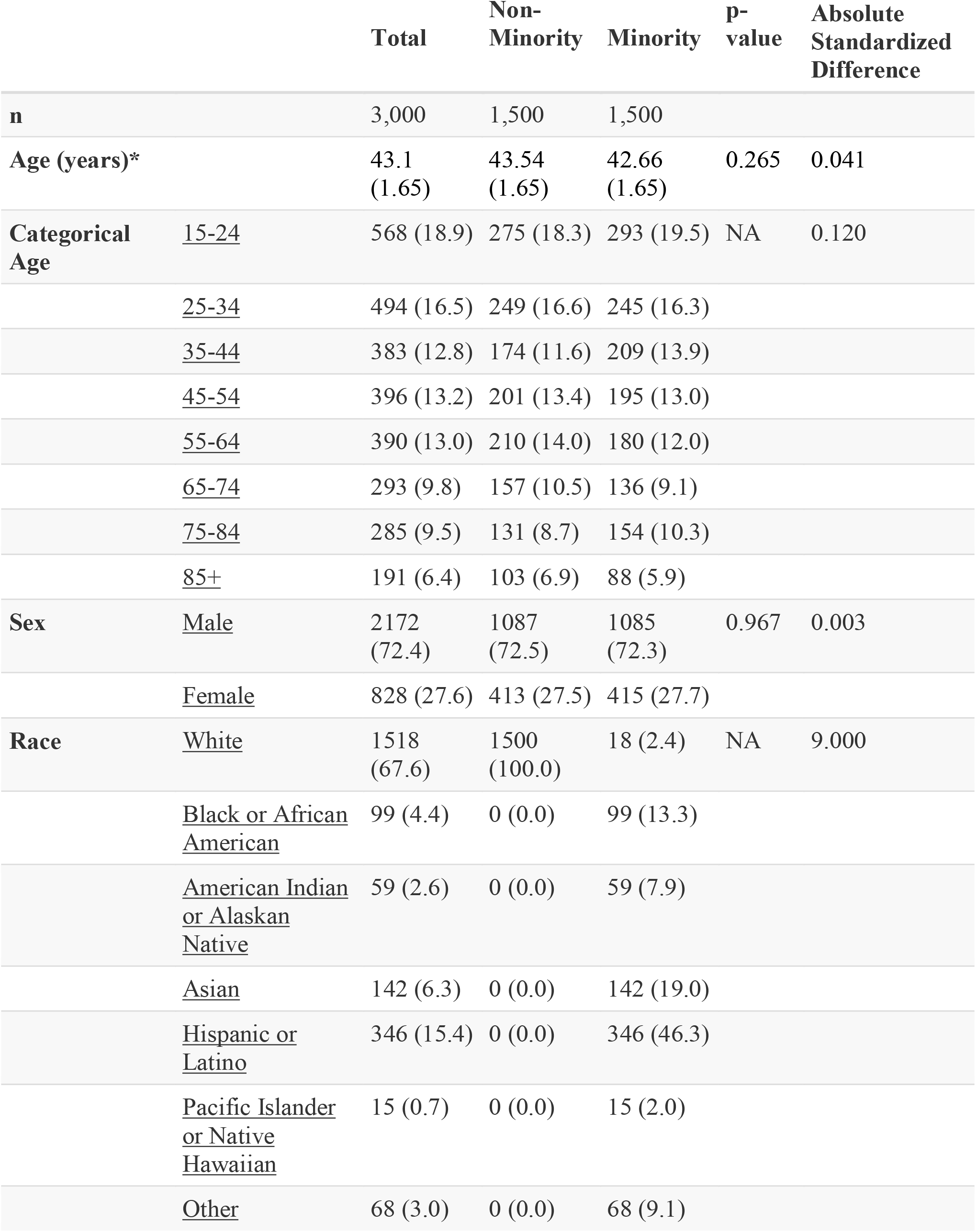
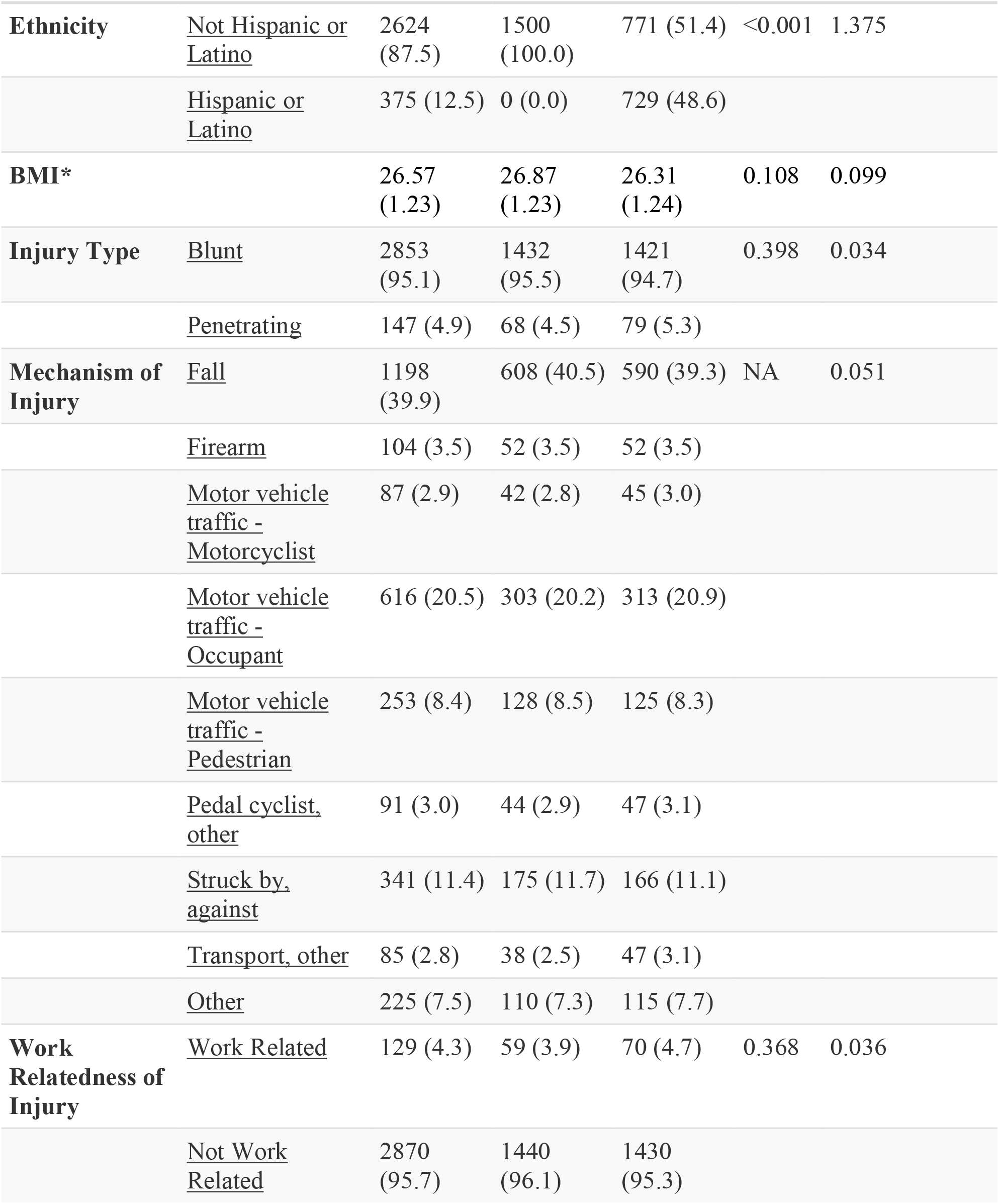
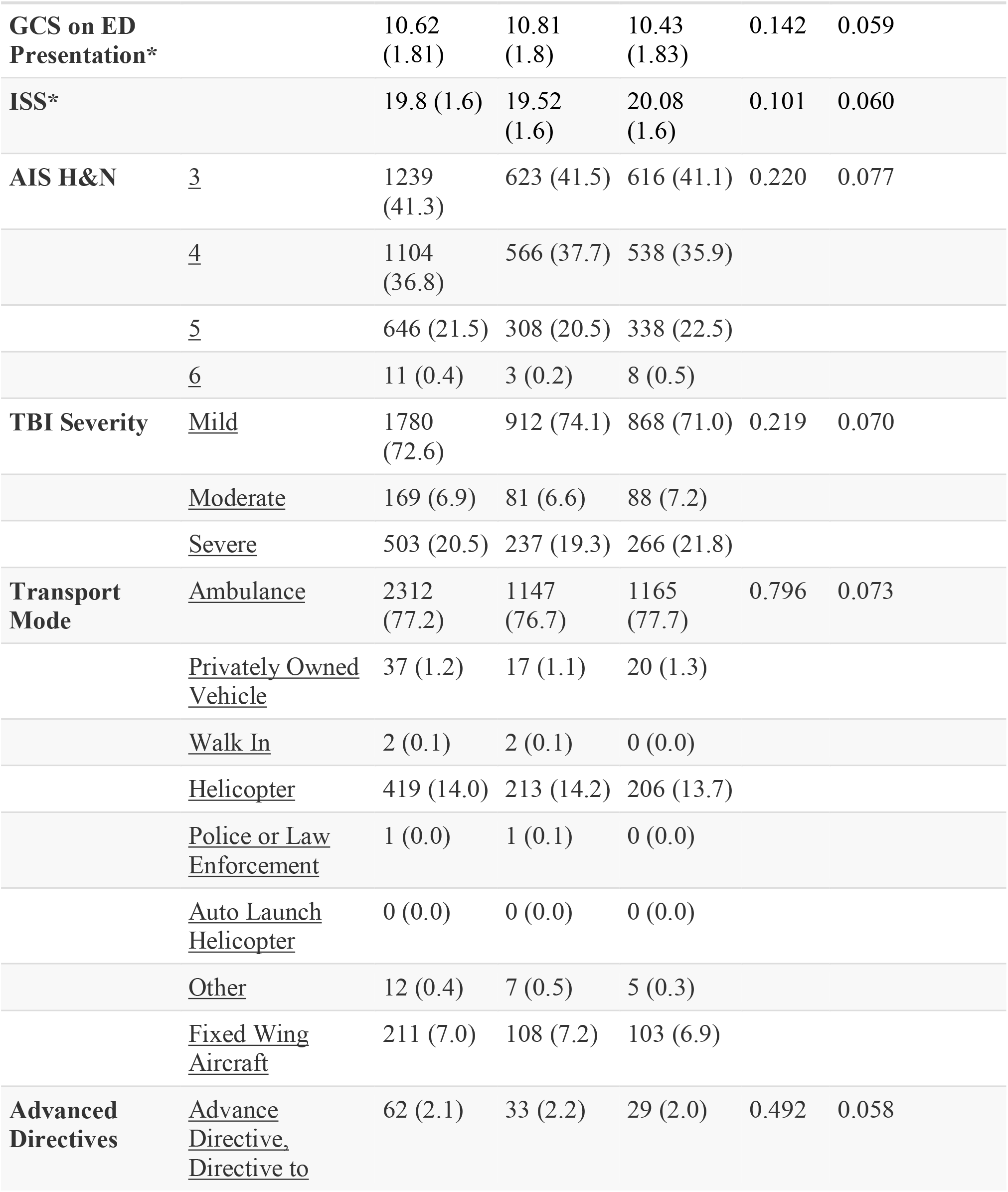
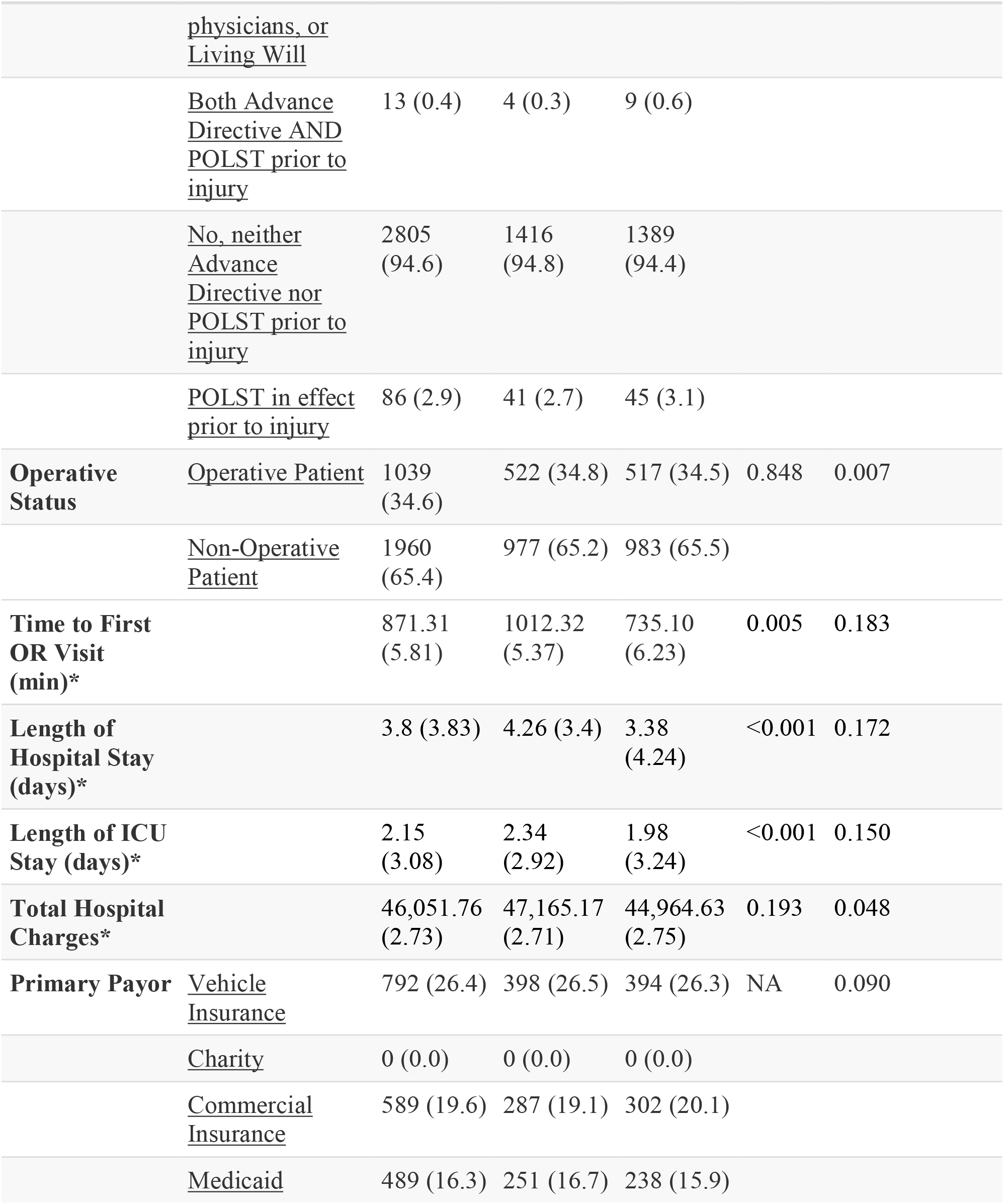
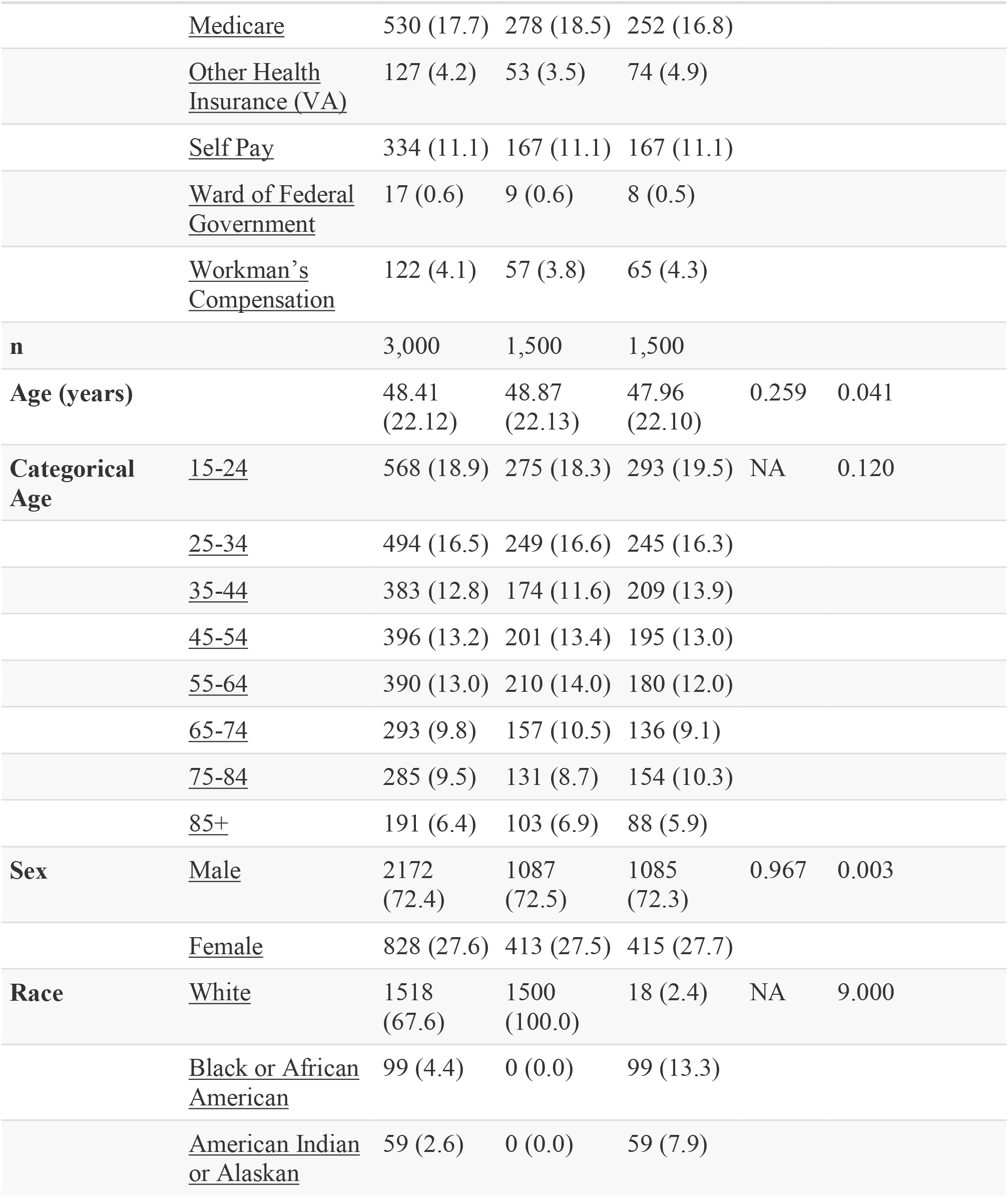
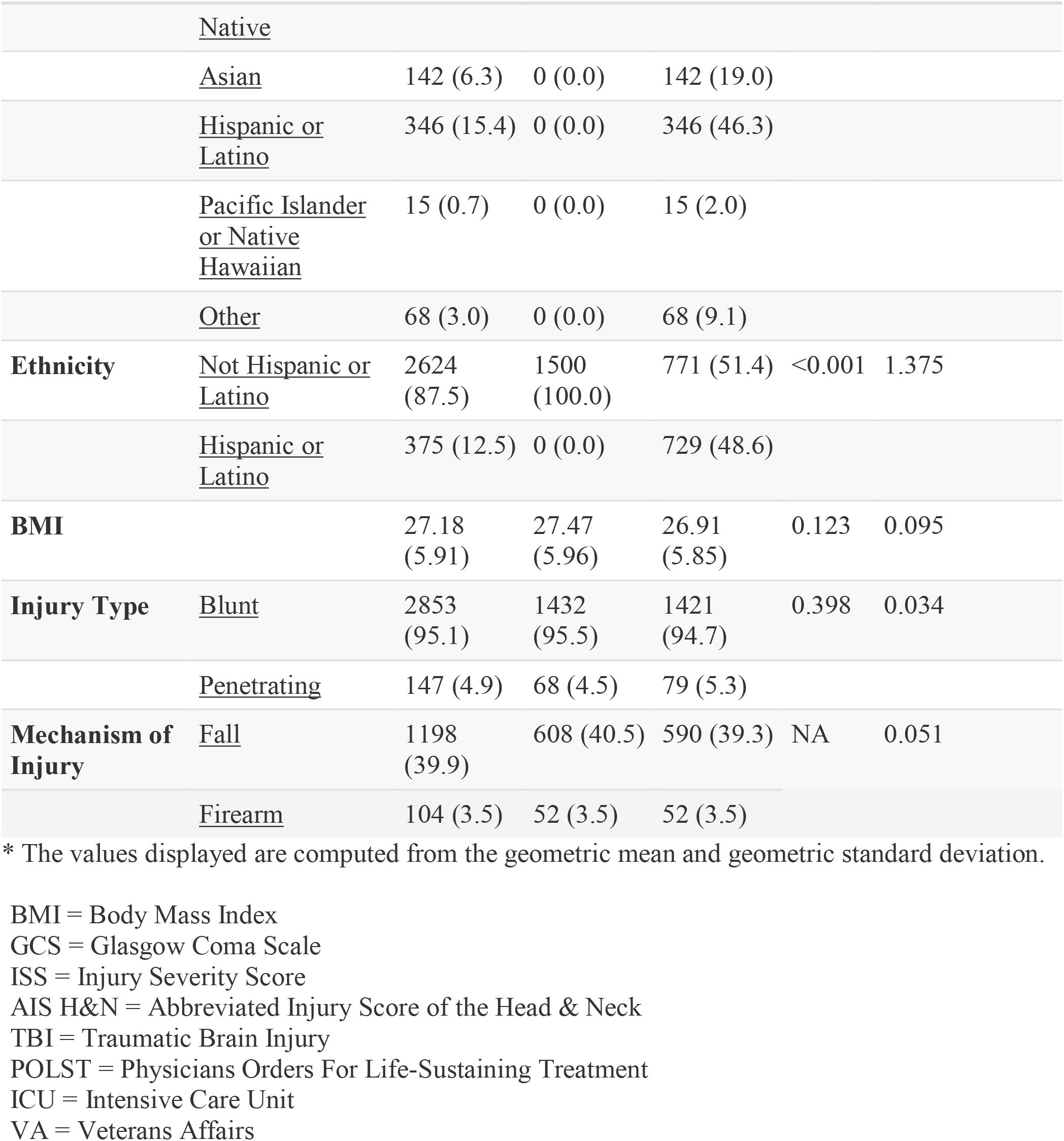
Patient characteristics stratified by minority status following propensity matching.

Overall, the patients were 68.5% male and 31.5% female. There was a statistically significant difference in sex between the White/non-Hispanic and the minority cohorts with males making up 67.3% of the White/non-Hispanic group vs. 72.4% of the minority cohort (p < 0.001). The average age of all patients was 53.25 years old. There was a statically significant difference in the average ages based on cohorts with that of the White/non-Hispanic group as 54.90 years old and of the minority group as 47.91 years old (p< 0.001). By age group, the highest incidence of TBI overall and in the White/non-Hispanic cohort occurred in the 55-64 year old group (14.9% of overall patients, 15.8 % of White/non-Hispanic patients) while the highest incidence for the minority cohort occurred in the 15-24 year old group (19.6% of minority patients).

Overall, there were 7 different categories for race/ethnicity included in the demographics of the database: White (86.9%), Black/African American (1.8%), American Indian or Alaskan Native (1.1%), Asian (2.5%), Hispanic/Latino (6.2%), Pacific Islander or Native Hawaiian (0.3%), and Other (1.2%). Of the minority sub-group, the patients were 13.4% Black, 7.9% American Indian/Alaskan Native, 19% Asian, 46.2% Hispanic/Latino, 2.0% Pacific Islander/Native Hawaiian, and 9.2% Other.

### 3.2 Mortality and adverse outcomes

The overall in-hospital mortality rate was 9.9% with a statistically significant difference between in-hospital mortality rate of White/non-Hispanic patients (8.5%) and the in-hospital mortality rate of minority patients (14.6%) (p < 0.001). The adjusted hazard ratio for in-hospital mortality for minority patients in the propensity-matched cohort was 2.21 (95% Confidence Interval (CI) 1.43, 3.43) with p < 0.001 (Table 2). Examination of the absolute standardized mean differences, and of the region of common support indicated effective propensity-score matching (Supplementary Figure 1, Supplementary Figure 2). There was no statistically significant difference in the overall probability of survival prior to propensity matching, however the Kaplan-Meier curve of both the non-propensity matched and propensity matched patients (Figure 1A, 1B) demonstrates a visibly lower survival probability in the minority group. The overall length of hospital stay was 7.70 days with a larger standard deviation in the minority group (12.08 days) vs the White/non-Hispanic group (10.65 days) (p = 0.435). The length of ICU stay in all groups was an average of 3.88 days (p = 0.966). There was no statistically significant difference in discharge disposition, with 58.5% overall discharged to home, 58.5% of White/non-Hispanic patients discharged home, and 60.0% of minority patients discharged home. There were slight differences in discharges to Skilled Nursing Facilities (SNFs) (17.9% of White/non-Hispanic patients vs 12.2% of minority patients), Acute Care Facility-Hospitals (1.6% of White/non-Hispanic patients vs 0.8% of minority patients), and Rehabilitation Facilities (5.2% of White/non-Hispanic patients vs 4.7% of minority patients).

**Table 2:** Final Cox Proportional Hazard regression models for overall survival using propensity-matched cohorts and robust standard error.

| **Mortality** |  | **n (%)** | **Univariate HR**<br>**(95% CI)** | **Multivariate HR**<br>**(95% CI)** |
| --- | --- | --- | --- | --- |
| Minority Status | <u>Non-Minority</u> | 1500<br>(50.0) | Reference | Reference |
|  | <u>Minority</u> | 1500<br>(50.0) | 1.83 (1.47-2.29,<br>p<0.001) | 2.21 (1.43-3.41,<br>p<0.001) |
| Injury Type | <u>Blunt</u> | 2853<br>(95.1) | Reference | Reference |
|  | <u>Penetrating</u> | 147 (4.9) | 3.50 (2.52-4.89,<br>p<0.001) | 1.51 (0.64-3.59, p=0.350) |
| Probability of Survival | <u>Mean (SD)</u> | 0.9 (0.1) | 0.02 (0.01-0.04,<br>p<0.001) | 0.04 (0.02-0.08,<br>p<0.001) |
| Operative Status | <u>Operative Patient</u> | 1039<br>(34.6) | Reference | Reference |
|  | <u>Non-Operative Patient</u> | 1960<br>(65.4) | 2.11 (1.66-2.69,<br>p<0.001) | 2.60 (1.55-4.35,<br>p<0.001) |

HR = Hazard Ratio
SD = Standard Deviation

**Figure 1A.**
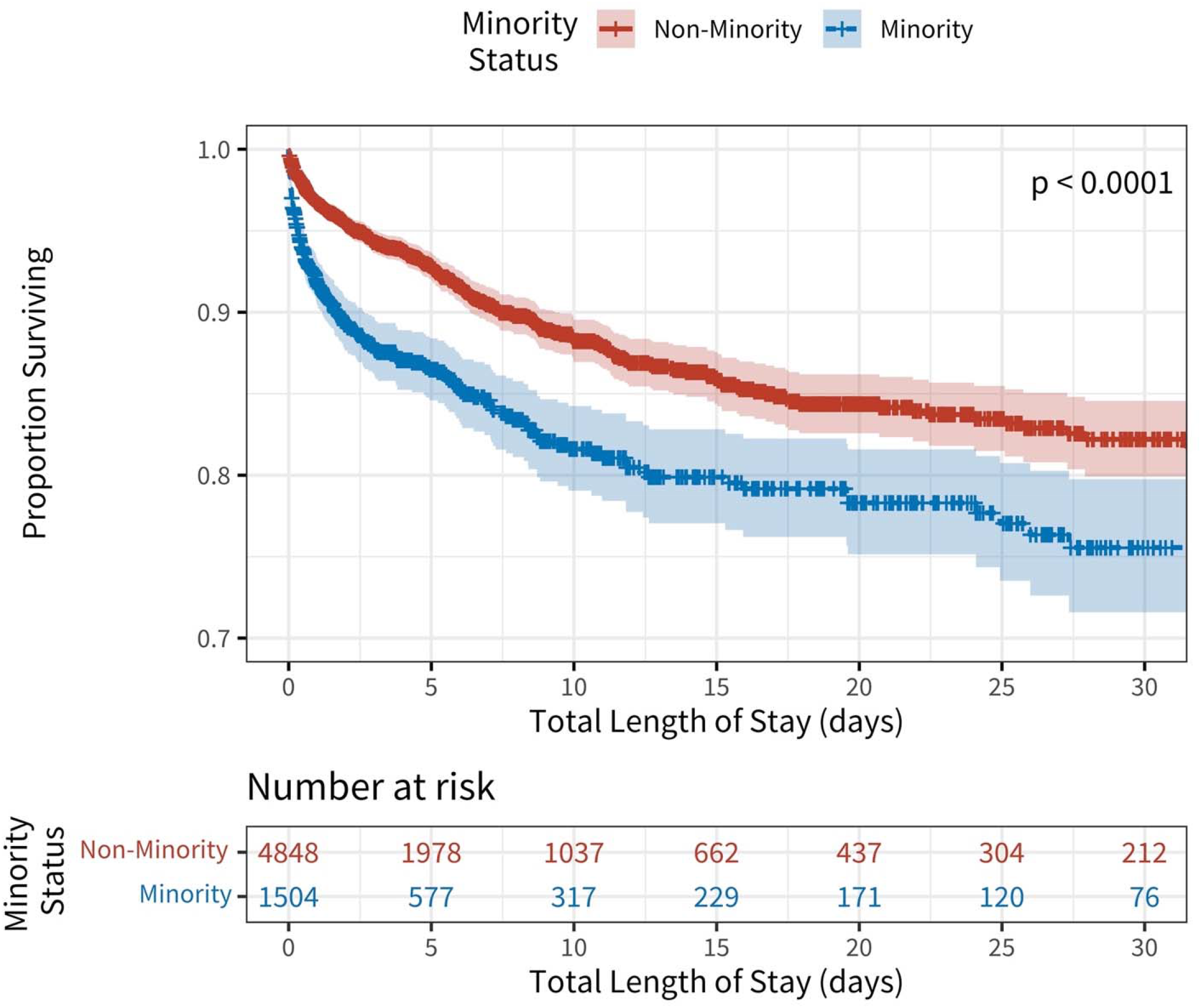
Unadjusted Kaplan-Meier survival curves stratified by minority status.

**Figure 1B.**
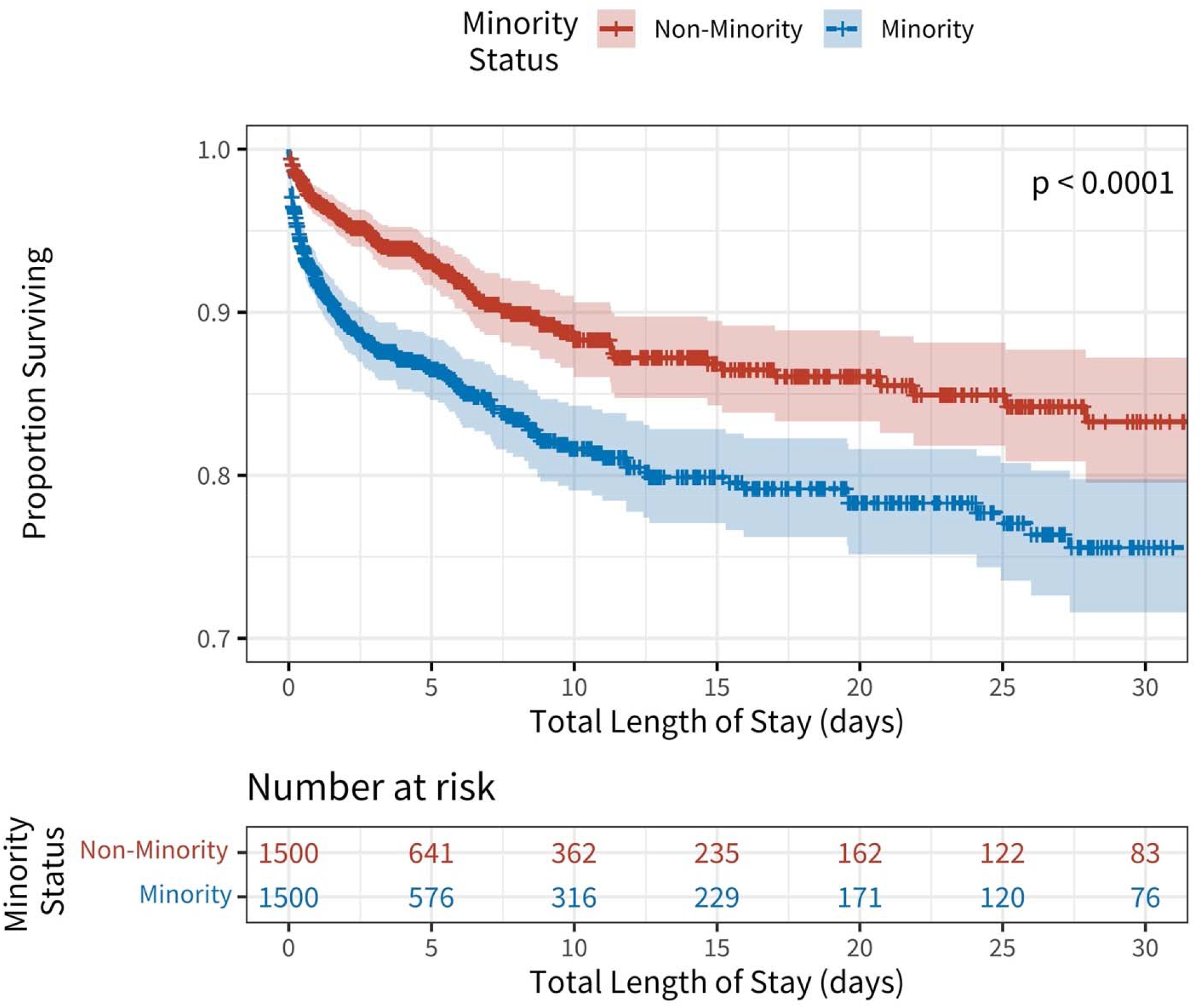
Propensity-Score Weighted Kaplan-Meier survival curves stratified by minority status.

### 3.3 Severity and type of disease at presentation

Out of all TBI admissions, 96.5% were due to blunt trauma and 3.5% were due to penetrating trauma. There was a statistically significant difference between percentage of blunt vs penetrating trauma with only 2.9% of trauma being penetrating in the White/non-Hispanic group while 5.3% of trauma was penetrating in the minority group (p < 0.001). In all patients, the most common mechanism of injury was fall (47.5%), followed by occupant in a motor vehicle crash (MVC) (17.0%). In the White/non-Hispanic group, this trend was similar with 50% of trauma being due to fall and 15.8% being due to MVC; however, in the minority group, although the trend was similar, the percentage of fall (39.3%) was lower and the percentage of MVC trauma (20.9%) was higher than the White/non-Hispanic group. There was a statistically significant difference in work-relatedness of injury with 4.7% of the minority group’s injuries being due to work vs only 3.2% of the White/non-Hispanic group (p = 0.011).

There were many statistically significant differences in severity of presenting injury between the White/non-Hispanic group and the minority group. The presenting GCS in the White/non-Hispanic group was 12.60 while that in the minority group was 11.95 (p < 0.001). The ISS in the White/non-Hispanic group was 21.27 vs 22.44 in the minority group (p < 0.001). The percentage of patients with AIS > 3 and 4 was similar between groups, however the percentage of patients with AIS > 5 was 18.5% in the White/non-Hispanic group vs 22.5% in the minority group. The percentage of TBI severity (mild, moderate, and severe) was statistically significant with 5.8% moderate and 16.4% severe TBI in the White/non-Hispanic group vs 7.2% moderate and 21.7% severe in the minority group (p < 0.001).

### 3.4 Types of medical interventions

There was no statistically significant difference between groups regarding mode of transport to the hospital with each group arriving via ambulance around 77% of the time and a privately owned vehicle 1.3-1.4% of the time. Advanced directives were in place for 3.0% of White/non-Hispanic patients vs 2.0% of minority patients. There were 92.5% of patients in the White/non-Hispanic group with neither an Advanced Director nor POLST in place prior to injury vs 94.4% of patients in the minority group with neither document in place. In the White/non-Hispanic group, 30.8% of the patients were operative vs 34.5% of the patients in the minority group (p = 0.008). The time from presentation to OR for White/non-Hispanic patients was 1048.08 minutes vs 735.71 for minority patients (p<0.001) (Table 3).

### 3.5 Insurance and financial factors

The average total hospital charges for all patients was $75,966.76 vs $75,408.30 for White/non-Hispanic patients and $77,766.92 for minority patients (p = 0.450). For the White/non-Hispanic group, commercial insurance was the most common primary payor at 31.5% of patients while for the minority group, the most common primary payor was vehicle insurance at 26.2% of patients. The second most common primary payor for White/non-Hispanic patients was Medicare at 21.4% while the second most common for minority patients was commercial insurance at 20.2%. Workman’s compensation paid as the primary payor for 2.9% of White/non-Hispanic patients vs 4.3% of minority patients. White/non-Hispanic patients utilized self-pay methods 7.3% of the time vs 11.1% of the time for minority patients.

## 4 Discussion

In this study, we explored the disparities between white/non-Hispanic and minority patients in mortality and adverse outcomes in traumatic brain injury patients presenting to a Level I trauma center in Portland, OR. Our results suggest that there are significant differences in mortality and adverse outcomes even given similar severity of injury, presenting factors, and financial factors. In our primary outcome, mortality, there was a large statistically significant difference between in-hospital death and hospital length of stay between groups. Accounting for potentially confounding factors such as severity of presenting injury, primary payor insurance mechanisms, hospital charges, and mechanism of injury, minority patients were at double the risk of in-hospital mortality than White/non-Hispanic patients. Regarding disparities in other aspects of care, there was no statistically significant difference in discharge disposition, though there was a trend towards higher rates of discharge to SNFs, Acute Care Facility-Hospitals, and Rehabilitation Facilities in the White/non-Hispanic group compared to the minority group. This could potentially demonstrate differences in access to such facilities, differences in insurance or payment methods, or even access to social work facilitation of these types of discharges.

Although there was no statistically significant difference in total hospital charges, our analyses did in fact show a trend regarding types of primary payors between the White/non-Hispanic patients and the minority patients in that the most common primary payor for White/non-Hispanic patients was commercial insurance vs vehicle insurance for minority patients. This correlates well to our finding that there was a higher percentage of MVC-related injury in the minority group compared to the White/non-Hispanic group and may also help to explain their greater severity of injury at presentation. However, the additional increase in percentage of both Workman’s compensation as well as self-pay seen in the minority group raises the concern that through these payor routes, minority patients may not have had similar access to discharge options such as SNFs or rehab facilities as did patients in the White/non-Hispanic group. Previous research has suggested that the influence of insurance is so strong that differences in coverage can increase the likelihood of availability to post-hospitalization care in less vulnerable populations^(30)^. Yet later research has explored the extent of influence insurance has on discharge location and has found that despite similar insurance, minority patients were found to be discharged more frequently to home without care than to SNF or acute-care facility post-hospitalization. Similarly, these studies found that white patients with lower vulnerable group membership (VGM) were discharged quicker to SNF or acute-rehab facility, which has been shown to provide higher rates of functional recovery than home care, than patients of color with higher VGM scores^(31-33)^.

There was a statistically significant difference in types of trauma between White/non-Hispanic patients and minority patients with minority patients presenting more often with penetrating trauma than their White/non-Hispanic counterparts. Minority patients also tended to have a higher rate of MVC as primary MOI and a higher rate of injury related to their work compared to the White/non-Hispanic patients. Minority patients tended to present with more severe injuries as demonstrated by both pure ISS as well as their tendency towards higher rates of more severe AIS at presentation. This may correlate to our findings that a higher number of minority patients were judged as operative patients compared to the White/non-Hispanic patients and similarly that minority patients had quicker times to OR than White/non-Hispanic patients. Although all of these findings as secondary outcomes are disparities in and of themselves, they do not explain the significant difference in our primary outcome of overall mortality between minority and White/non-Hispanic patients given that our primary analysis accounted for these severity factors.

It is well-studied in many other aspects of medicine and epidemiology that patients receive differing quality of healthcare and therefore have significantly different outcomes as a result of these health disparities. Not only do minority patients face persistent unconscious bias in the healthcare field and have differential access to baseline healthcare, it has also been shown that the healthcare that they do end up receiving likely is less adequate to handle surgically or medically complex issues due to shortages of necessary specialists, technology gaps, or timely access to services^(24, 34)^. Previous research in neurosurgery and other surgically complex fields have demonstrated marked disparities in the procedural outcomes between minority and White/non-Hispanic patients^(13, 22, 23, 35)^, however the current research regarding disparities in trauma management of minority patients still has yet to show the true inequality of care.

Among reasons that can be documented for these healthcare disparities in traumatic brain injury, one of the most likely explanations in our patient population was the sheer severity of injury at presentation and the likelihood of operation in our minority patients. This, alongside greater numbers of penetrative traumas and traumas related to MVC rather than fall, produced a much more severe clinical picture for our minority group than our White/non-Hispanic group. To explain these presenting findings, it is important to look at the history of trauma in regions and neighborhoods of predominantly minority persons. Through decades of housing red-lining, tax adjustments, and inequitable educational progress, neighborhoods primarily composed of minority persons in America have trended towards more dangerous, violent regions by no fault of the persons living in those regions^(36-40)^. It is logical to assume that areas of increased risk of violence would have higher rates of violent trauma than those with better protections in place. Additionally, persons of minority status often live further from healthcare facilities adequately equipped to manage the complexity of the trauma with which the patient presents. Many studies have recently shown the extent of the disparities in both number and quality of providers but also the number and quality of the technology available at local or regional hospitals compared to the wealthier and often more academic hospitals placed in centers of higher economic advantage^(12, 24, 34, 41).^

However, there are many factors that could potentially influence a patient’s mortality or secondary outcomes following a traumatic brain injury. The limitations of this paper lie in the fact that many of these factors are simply not recorded in our typical point of care healthcare system for trauma and therefore are much more difficult to study. Additionally, the retrospective nature of this study is a limitation. However, most epidemiological and sociological research looking at cultural and community-oriented factors related to healthcare can only be studied in a retrospective manner. In this paper, we were not able to examine all the pre-hospital factors influencing our patients outcomes and thus could only analyze the data points associated with the seemingly extensive Trauma One database for one of the two level 1 trauma centers in the state of Oregon, yet even with these data, there are numerous sociological, epidemiological, financial, psychological, and political factors at play that go unreported and therefore are under-researched^(21)^. These factors typically thought of to be non-medical have consistently demonstrated pervasive, persistent, and poignant disparities between minority patients and White/non-Hispanic patients that have plagued our healthcare system in the United States^(1, 3, 8, 10, 38, 42-44)^. These health disparities have been demonstrated to be independent of individual health choices; these inequalities are not biologically determined yet they plague only certain sects of our populations, permeating beyond healthcare. Not only do our minority patients have generations of well-documented medical trauma that prohibits a full trusting relationship of the American healthcare system but there are clearly also disparities in patient’s lives pre-hospital such as access to routine healthcare, leniency in the workplace for regular healthcare maintenance, financial freedom for healthcare maintenance, safe working environments, housing opportunities in neighborhoods free of violence, full of healthy foods, and access to clear air/water, and many more that are simply not documented when our patients present with neuro-trauma^(3, 8, 38, 42, 44)^. The health equity framework with which we claim to utilize to progress our medical care demands that we recognize these extraneous but highly influential aspects of our patients’ lives. We must make ourselves aware of the macro-social factors which perpetuate this disproportionate suffering among our patients of color and act with the responsibility and privilege we carry in our communities to make these disparities known. We must not only care for our patients when we encounter them within our clinics, emergency departments (EDs), and operating rooms, but in the healthcare disparities that exist outside of the concrete confines of the traditional healthcare system. Thus, we have a responsibility to engage with the upstream factors in our patients’ lives that put them at increased risk even prior to entering our facility’s doors. The traumas of structural racism do not start the moment of impact from a fall or an MVC; our patients’ traumas begin when they are raised in a discriminatory society and robbed of certain medical, social, political, and financial privileges accustomed by our White/non-Hispanic patients.

Our study is but one current research set to analyze the disparities in yet another aspect of medical care in the United States and certainly not the only one to find inequalities in management of trauma. It is imperative that we as a medical community target not only healthcare improvements to properly regulate and treat neurosurgical trauma, but focus our progress towards equitable community environments, living situations, financial and insurance-based payment plans, and trauma prevention efforts in order to better care for our patients of color both pre-hospital and once they arrive into our ED. Now with an ever-growing accumulation of research shining light to these racial and ethnic disparities in every corner of medical research, it is simply insufficient to produce more research, but it is ever more pertinent to call all medical providers to greater action.

## 5 Conclusion

In this study, we explored the in-hospital mortality outcomes and characteristics of adult patients who were admitted with traumatic brain injury to one of the two Level I trauma centers in Portland, OR between 2006 to October of 2017. Our data demonstrate a significant disparity in overall in-hospital mortality in minority patients, greater severity of presenting injury, higher incidence of MVC- or work-related injury, and higher operative rate than White/non-Hispanic patients given insignificant differences in insurance, hospital charges, and other accounting factors. Although these are novel findings for the field of neurosurgical trauma related to traumatic brain injury, they are repeated in other specialties and other medically complex patient populations in the American healthcare system. Research has been keen to identify these marked inequalities over the past couple of decades yet still there has not been a clear path to dismantling these disparities within the medical community. Treating patients under our care is our greatest privilege and responsibility as physicians. As such, we have a societal and professional duty to recognize and accept that the effects of structural racism have taken hold of our patients’ health long before they arrive in our trauma bays, ICU beds, and operating tables. These disparities permeate our society and contribute to inequitable health outcomes, and we must take action to identify the factors which perpetuate this disproportionate suffering. Simply treating the minority of patients who require surgical intervention or clinical consultation is not enough. Our roles demand that we recognize these larger social factors acting upstream on our patients before they enter our fractioned healthcare system which often fosters the very mistrust that hides them from our otherwise watchful eyes in the first place. The necessary first step in providing an equitable healthcare system for all patients regardless of socioeconomic, racial, or ethnic status, is continued awareness of the pervasiveness and consistency of these medical disparities and emphasized efforts to improve the sociological factors that perpetuate them.

## Data Availability

The original contributions presented in the study are included in the article/supplementary material, further inquiries can be directed to the corresponding author/s.

