## Supplementary Figures for "Racial and Ethnic Inequities in Mortality During Hospitalization for Traumatic Brain Injury: A Call to Action" (from another version of this preprint: version 1, 2021-04-09)

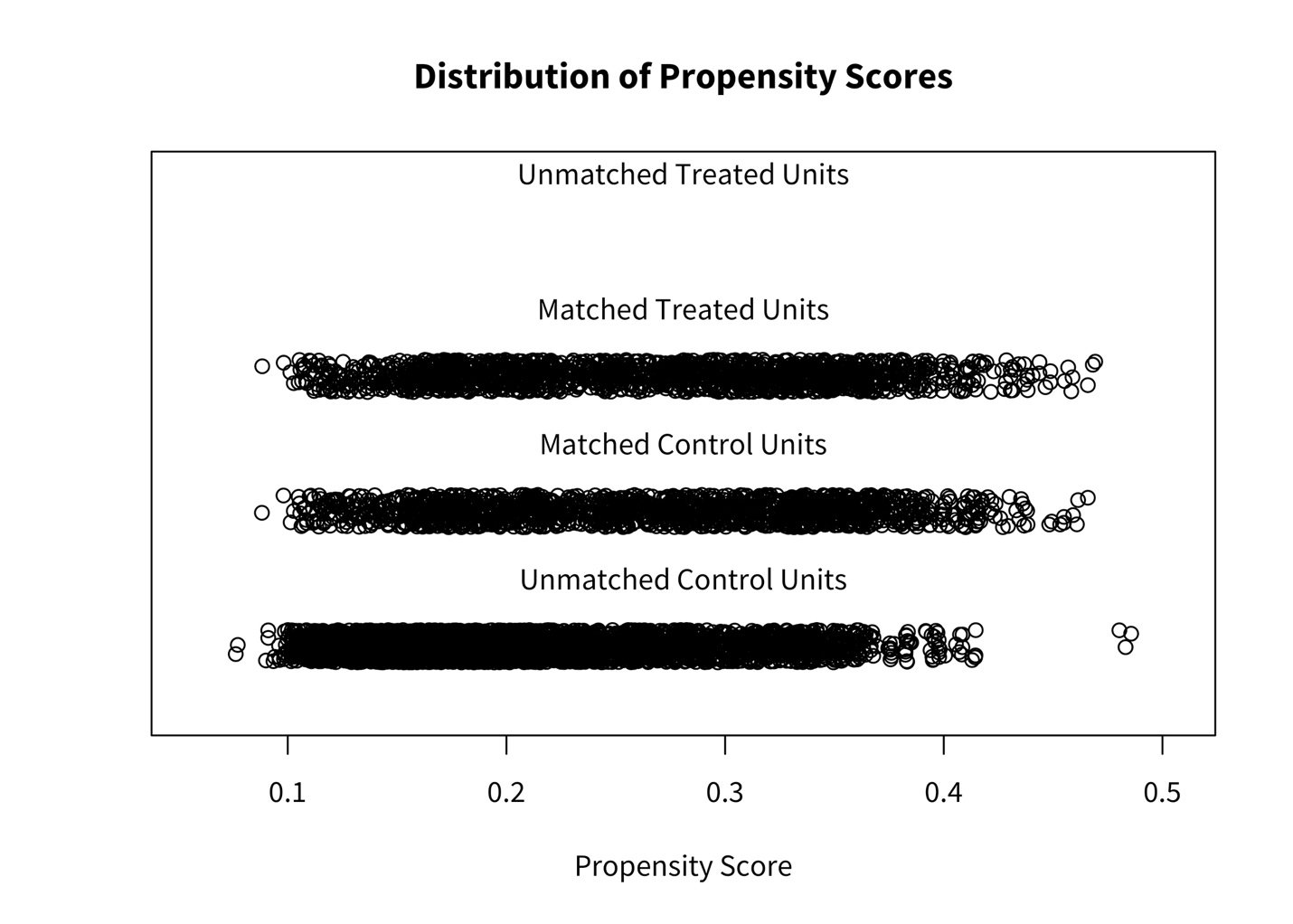

**Supplementary Figure 1.** Jitter plot of distribution of propensity scores before and after matching, including unmatched controls.

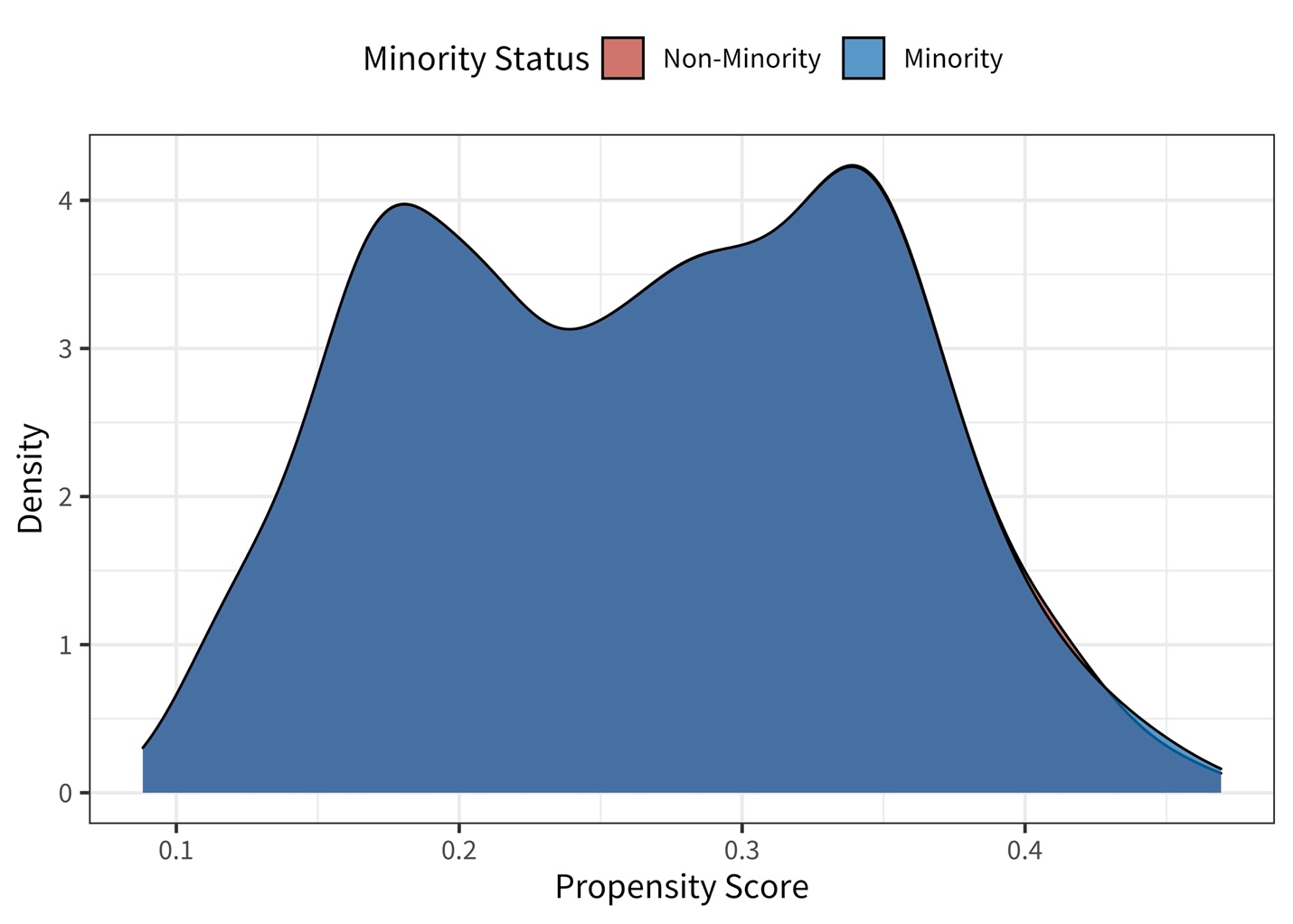

**Supplementary Figure 2.** Histogram of density of propensity score distribution demonstrating region of common support.

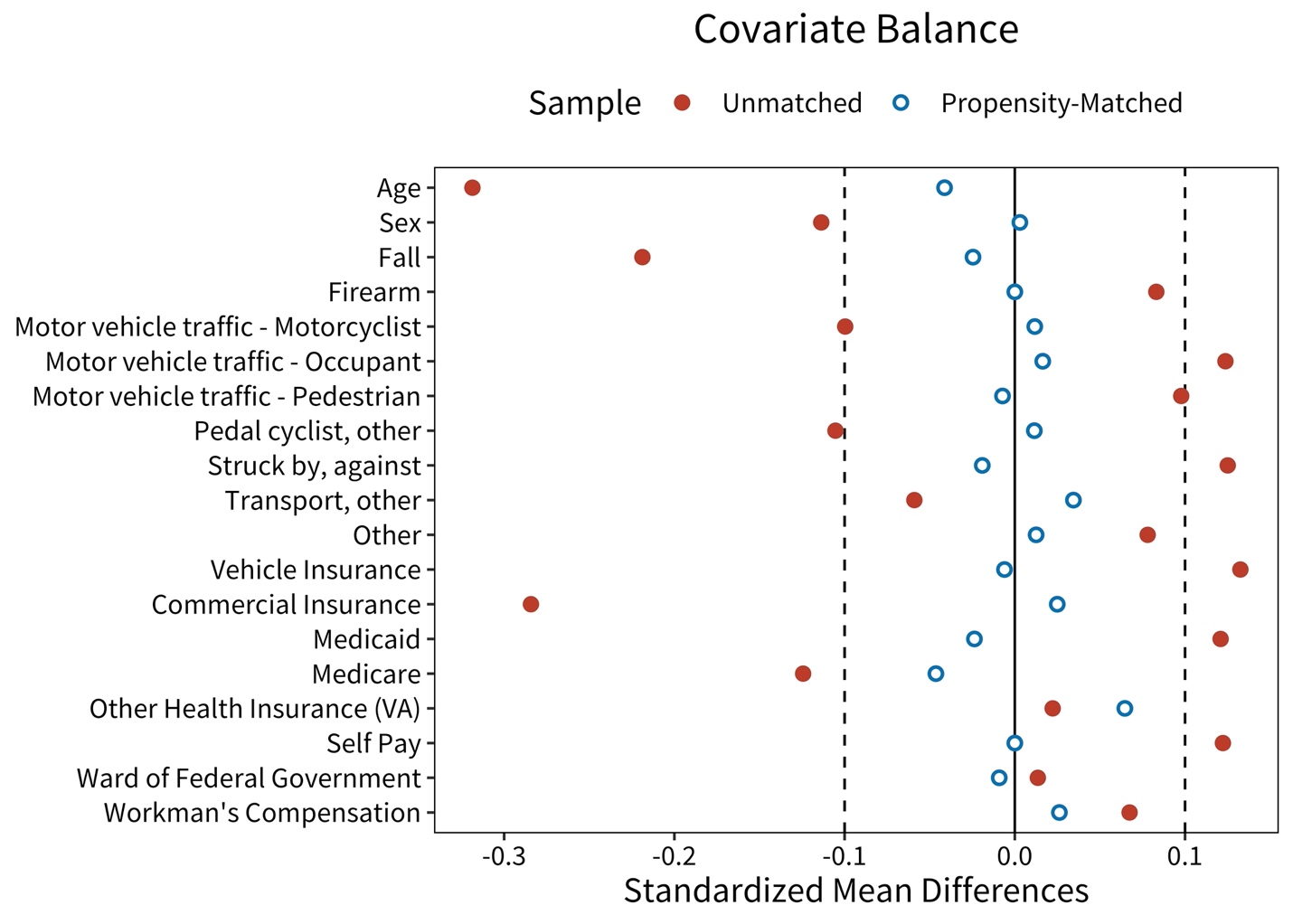

**Supplementary Figure 3.** Graphical comparison of standardized mean differences assessing covariate balance before and after propensity score matching.
